# Analysing and comparing the COVID-19 data: The closed cases of Hubei and South Korea, the dark March in Europe, the beginning of the outbreak in South America

**DOI:** 10.1101/2020.04.06.20055327

**Authors:** Stefano De Leo, Gabriel G. Maia, Leonardo Solidoro

## Abstract

The present work is a statistical analysis of the COVID-19 pandemic. As the number of cases worldwide overtakes one million, data reveals closed outbreaks in Hubei and South Korea, with a new slight increase in the number of infected people in the latter. Both of these countries have reached a plateau in the number of Total Confirmed Cases per Million (TCCpM) residents, suggesting a trend to be followed by other affected regions. Using Hubei’s data as a basis of analysis, we have studied the spreading rate of COVID-19 and modelled the epidemic center for 10 European countries. We have also given the final TCCpM curves for Italy and Lombardia. The introduction of the *α*-factor allows us to analyse the different stages of the outbreak, compare the European countries amongst each other, and, finally, to confront the initial phase of the disease between Europe and South America.

**Methods:** By dividing the TCCpM curves in multiple sections spanning short time frames we were able to fit each section to a linear model. By pairing then the angular coefficient (*α* factor) of each section to the total number of confirmed infections at the center of the corresponding time interval, we have analysed how the spreading rate of Covid-19 changes as more people are infected. Also, by modelling the TCCpM curves with an asymmetrical time integral of a Normal Distribution, we were able to study, by fitting progressively larger data ensembles, how the fitting parameters change as more data becomes available.

**Findings:** The data analysis shows that the spreading rate of COVID-19 increases similarly for all countries in its early stage, but changes as the number of TCCpM in each country grows. Regarding the modelling of the TCCpM curves, we have found that the fitting parameters oscillate with time before reaching constant values. The estimation of such values allows the determination of better parameters for the model, which in turn leads to more trustworthy forecasts on the pandemic development.

**Interpretation:** The analysis of the oscillating fitting parameters allows an early prediction of the TCC, epidemic center and standard deviation of the outbreak. The *α* factor and the recovered over confirmed cases ratio can be used to understand the pandemic development in each country and to compare the protective measures taken by local authorities and their impact on the spreading of the disease.

**Funding:** CNPq (grant number 2018/303911) and Fapesp (grant numebr 2019/06382-9).

## I. Introduction

The COVID-19 outbreak is caused by a newly identified member of a large family of viruses known as coronaviruses (name deriving from the Latin word *corona*, meaning crown). According to the World Health Organization (WHO) [1] this family of viruses causes illnesses ranging from the common cold to more severe diseases such as Severe Acute Respiratory Syndrome (SARS) and the Middle East Respiratory Syndrome (MERS). These viruses were originally transmitted from animals to people. SARS, first recognized in mid-March 2003 [2], was transmitted from civet cats to humans, while MERS, first detected in Saudi Arabia in 2012 [3], moved to humans from a type of camel. The novel coronavirus, identified by Chinese authorities on January 7 is a new strain that has not been previously identified in humans. The SARS-CoV-2 virus, as it has been named by the WHO, seems to be closely related to bat and pangolin coronaviruses, but a final confirmation has not yet been given [4]. People may catch COVID-19 by close contact with infected people, via small droplets produced when they cough, sneeze, talk, or by touching a contaminated surface and then their face. According to the WHO, symptoms of infection include fever, cough, shortness of breath and breathing difficulties. Notwithstanding the fact that infected patients can also be asymptomatic, in the more severe cases, the virus can lead to pneumonia, multiple organ failure and death. The number of fatalities from the new coronavirus has overwhelmingly overtaken the toll of the SARS outbreak but its percentage remains considerably lower compared to the 10 percent of SARS and 30 percent of MERS.

Chinese health authorities are still trying to determine the origin of the virus. By the end of 2019, the Chinese office of WHO received reports on an pneumonia of unknown cause in the city of Wuhan, located in the Hubei province. At the beginning of 2020, the cases developed into a cluster of pneumonia, which started being addressed by both WHO and local authorities. On January 13th, the first confirmed case outside of China, a patient in Thailand, showed the disease was no longer confined to the Chinese territory. In two months, on the 7th of March, the 100.000 cases mark was reached [5] and four days later WHO declared the COVID-19 a pandemic, with Europe being recognised as the new epicenter shortly after. As of the time of the writing of this paper, the beginning of April, nearly one million cases have been identified worldwide.

The present work analyses data on the daily development of COVID-19 cases in Hubei, South Korea, 10 European countries (Austria, Belgium, France, Germany, Italy, Netherlands, Portugal, the United Kingdom, Spain, and Switzerland), 4 Italian regions (Emilia-Romagna, Lombardia, Piemonte, and Veneto), and 10 South American countries (Argentina, Brazil, Bolivia, Chile, Colombia, Ecuador, Paraguay, Peru, Venezuela, and Uruguay). The data was collected by the websites [5–7] and the our statistical analysis begins on January 22th (day 1).

The paper is divided as follows: In the next section, we analyse the spreading rate of COVID-19 as a function of the confirmed cases. The TCCpM curves are not linear over long time intervals (weeks and months), but sections of it spanned over shorter periods can be approximated so. In this paper, we have chosen an interval of 5 days. By studying how the angular coefficient of these straight (called for simplicity of presentation the *α* factor) changes with the number of confirmed people, we have an insight on how COVID-19 spreads over the different European countries for a given TCCpM value. In section 3, we numerically fit progressively increasing portions of the available data on the TCCpM to a mathematical model, checking how our predictions of the model’s fitting parameters change with time as more data becomes available. The values of the parameters show an oscillatory behaviour which eventually reaches stability. The stable values can then be used in the model to forecast the development of the pandemic itself. Section 4 looks more closely at the Italian case by studying how the virus spread over 4 different Italian regions. Section 5 analyses the beginning of the outbreak in South America. In the final Section, we draw our conclusions.

## II. Spreading rate and the *α* factor

Fig. 1 shows the total number of confirmed cases per million residents in 10 European countries, the Chinese Province of Hubei and South Korea since day 0 (21 January) of the outbreak data. We can see that for nearly 30 days the spreading of COVID-19 is more or less confined to China, with a steep increase everyday. Around day 20 (10 February), the total number of confirmed cases in Hubei starts to stabilise to nearly 1200 TCCpM. A similar curve can be observed for South Korea. South Korea’s population is of comparable size to Hubei’s population, as can be seen in the table annexed to Fig. 1, but their TCCpM curve stabilises much faster and at a much lower value, approximately 200 TCCpM. As of now, no other country studied in this work has reached a similar stabilisation. This striking difference might be due to the timely action of the South Korean authorities in adopting preventive measures to contain the outbreak, including, among others, the large number of tests performed. For example at day 48 (9 March), when both Italy, with its 9172 total confirmed cases and South Korea, with its 7478 ones, reached 150 TCCpM, see Fig. 2, the number of tests performed in Italy (60761) was three and a half times smaller than the ones carried out in South Korea (210144).

**Figure 1:**
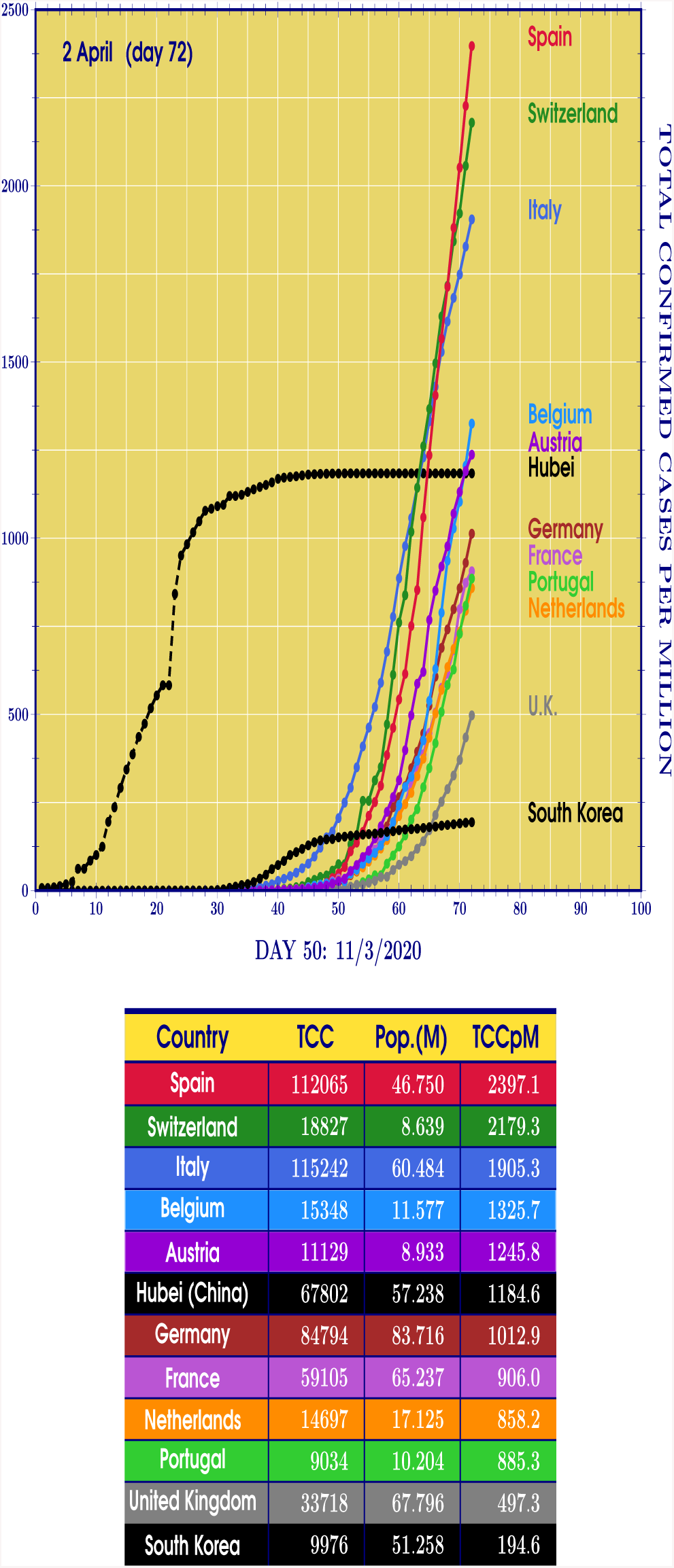
EUROPE, HUBEI, SOUTH KOREA. Curves of the Total Confirmed Cases per Million (TCCpM) of residents for Spain, Switzerland, Italy, Belgium, Austria, Italy, Germany, France, Netherlands, Portugal, the United Kingdom, and South Korea at the 2nd of April (day 72). At the beginning of the disease, we found, for all the countries, a slow increase of the curves. After the initial period, we see a steep increase in TCCpM. For Hubei and South Korea, we also observe that the curves reach a stabilisation point after a while, indicating the decreasing number of new cases per day. The plots show that this stabilisation point has not yet been reached by the European countries at day 72. The possibility to compare the different TCCpM increasing rates for each country is one of the main subject matter of our investigation.

**Figure 2:**
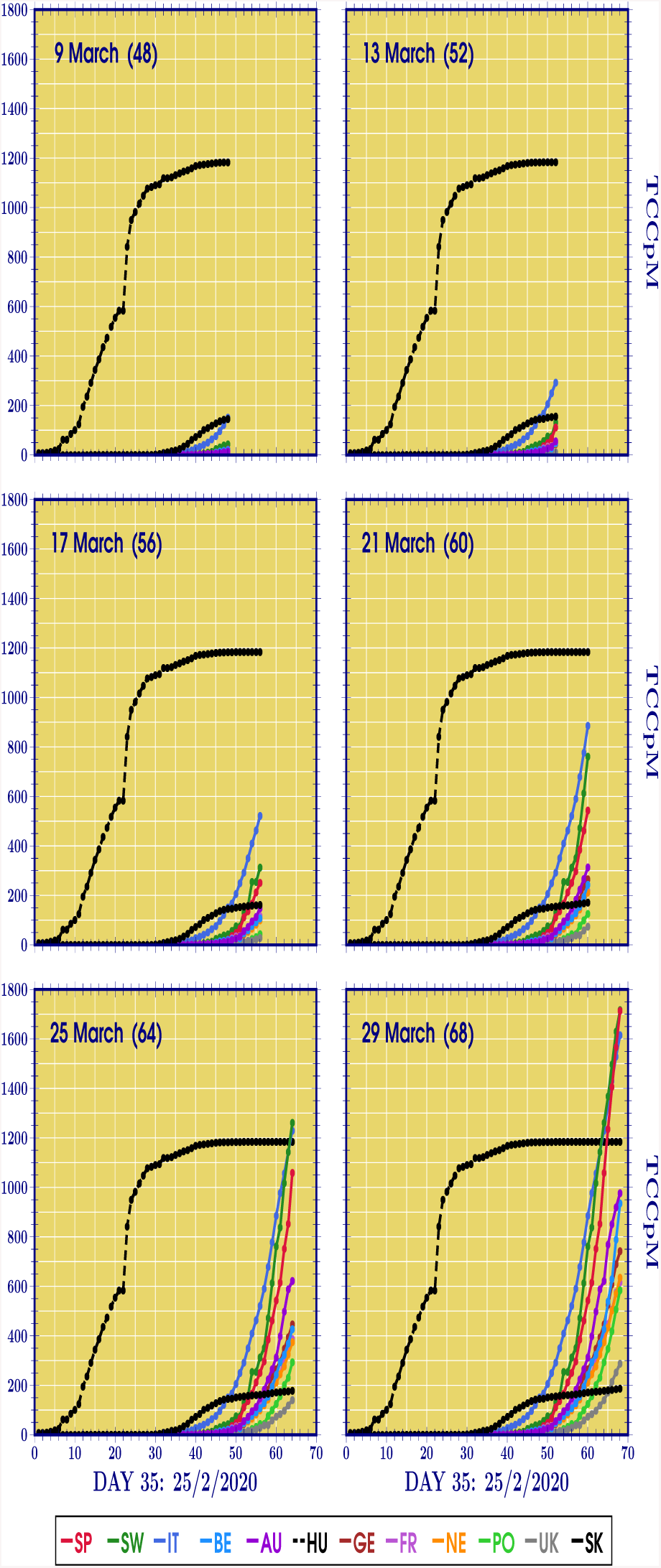
EUROPE,HUBEI, SOUTH KOREA. The time series of the TCCpM for the European countries of Fig. 1, Hubei, and South Korea are shown for different days. At day 48, Italy has the same TCCpM of South Korea, at day 52, Italy overtakes and Switzerland reaches South Korea, at day 60, many of the 10 European countries curves are located between the South Korea and Hubei ones, at day 64, Switzerland and Italy overtake Hubei, and, finally, after further 4 days, Spain overtakes Hubei and Italy and reaches Switzerland (overtaking it at day 72, see Fig. 1). The updated animated version of this figures is found in the website [8].

Fig. 2 gives a better idea of how the TCCpM spreading rate in the European countries changes from around day 50 up to around day 70. The time series show, for instance, that at day 48, Italy has the same TCCpM as South Korea. At day 52, Italy overtakes and Switzerland reaches South Korea. At day 64, Switzerland and Italy both overtake Hubei and, finally, after 4 days, Spain overtakes Hubei and Italy, and reaches Switzerland (overtaking it at day 72 as shown in Fig. 1). The complete time series can be found updated and in animated format in the website [8].

In order to introduce a simple parameter which allows comparisons between countries, let us look again at Fig. 1. The TCCpM curves do not behave linearly for any region until they stabilise. If, however, we look at each curve closely, in a span of only a few days the development of the number of cases can be approximated to a straight line. By fitting then the TCCpM for a period of five days to a linear model,

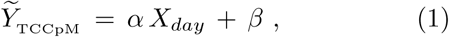

we find the angular coefficient *alpha* for each country for that time frame. This give us information about how fast COVID-19 spreads with time, consequently allowing us compare the spreading rate for different countries. The *β* term is the number of infected people at the beginning of the time span covered by the model. What we did then was to select five consecutive days and compute the TCCpM of each, fitting these data to a linear model from which we extract the *α*-factor. After this we pair this *α*-factor to the TCCpM calculated at the center of the time period and then move to another ensemble of five days and repeat the process. This provides us with a correlation for how fast COVID-19 spreads as more people are infected.

In Fig. 3(a) the *α*-factor is plotted for the 10 European countries, Hubei, and South Korea as a function of the TCCpM. The behaviour of the Hubei and South Korea’s curves, clearly shows that the *α* factor after reaching its maximum value goes to zero, closing the outbreak. It is also indicative that for South Korea, where the disease was soon taken at its beginning and the measures adopted timely, we found only a maximum (reproducing the well know Gaussian behaviour) while for the Hubei we find two local maxima. The double peak phenomenon is also found for the European countries. After an initial increase, the European curves start to develop in a more horizontal fashion, indicating the infection rate to approach a more linear growth. This horizontal development is not that of a constant, obviously, which would firmly indicate a linear growth, but it reveals slowing down of the spreading rate with the number of confirmed infections. For Italy, the *α* factor is in its decreasing region after the double peak. Fig. 3(b) allows a more detailed look at the *α*-factor from 0 to 500 TCCpM. At 500 TCCpM Switzerland starts to increase substantially compared to the other European countries. Let us understand the importance of this fact by comparing Switzerland with Italy. At the day 55 (16 March), Italy (with 27980 confirmed cases) has a TCCpM of 462.6 with an *α* factor of 59.3. Italy in that moment was adopting very restrictive preventing measures. At the day 58 (19 March) Switzerland (with 4075 confirmed cases) reaches 471.7 of TCCpM with an *α* factor of 116.0 almost twice greater.

**Figure 3:**
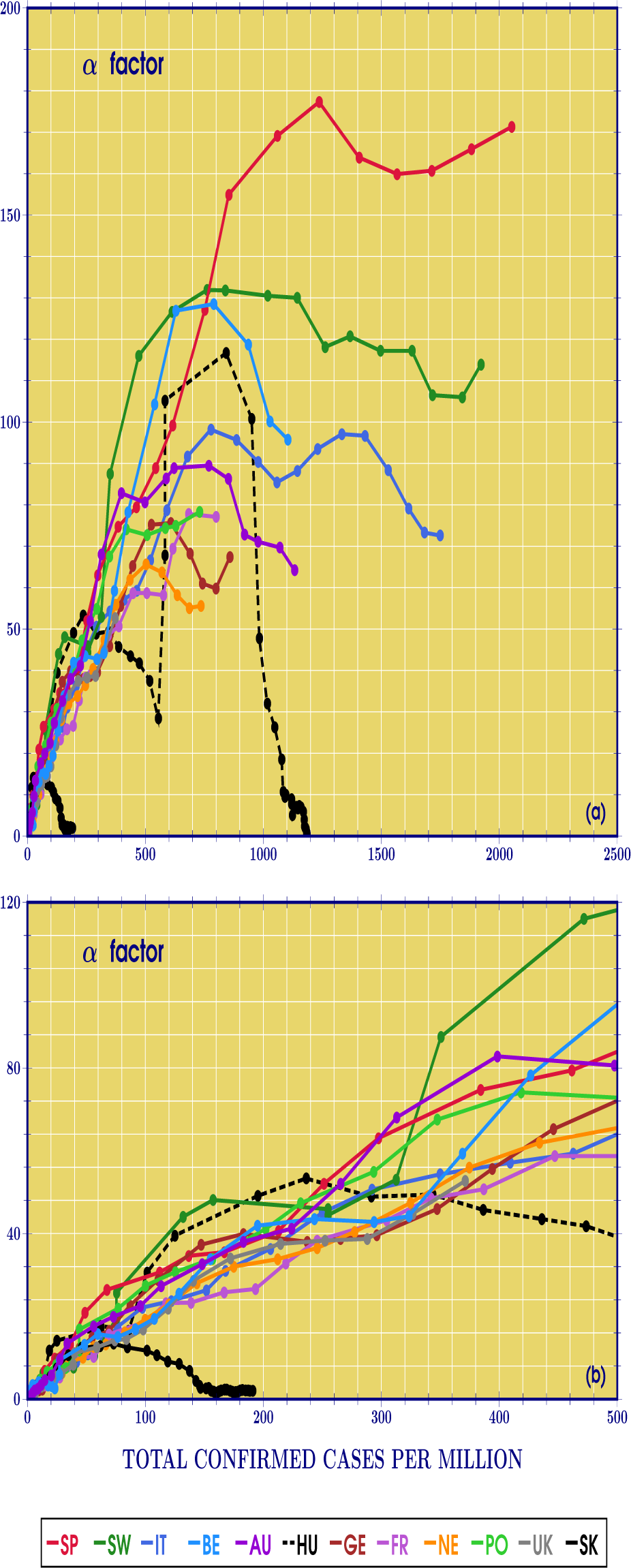
EUROPE,HUBEI, SOUTH KOREA. The *α*-factor, representing the spreading rate of COVID-19, is plotted in (a) for Spain (SP), Switzerland (SW), Italy (IT), Belgium (BE), Austria (AU), Hubei (HU), Germany (GE), France (FR), Netherlands (NE), Portugal (PO), the United Kingdom (UK), and South Korea (SK), as a function of the TCCpM. The bottom figure (b) allows a more detailed look at the *α*-factor from 0 to 500 TCCpM. We can see that, in the range from 0 to 300, the contagion rate for the European countries is approximately the same. Reaching 500 TCCpM, it becomes evident that Switzerland and Belgium start to increase substantially compared to the other European countries.

We conclude this Section by observing that a three days period of analysis, instead of the five used in our simulations, produces curves similar to the ones of the daily confirmed cases with a greater number of loops to be performed to complete the analysis. A week period has the advantage of a minor number of loops in the simulation but clearly does not represent a good approximation of the TCCpM curve.

The *α* factor is an interesting parameter to compare the spreading rate of different countries. In the next section, it will be complemented by other ones.

## III. Modelling the TCCpM curve

Taking Hubei’s TCCpM curve as an example of a COVID-19 pandemic development from its earliest stages to its conclusion, see Fig. 1, we model it by the following function

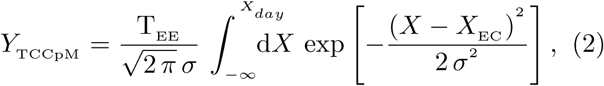

where T_EE_ is the TCCpM at the end of the outbreak, *X*_EC_ the Epidemic Center, and *σ* is the Standard Deviation [9].

Using the software *Wolfram Mathematica* [10], we applied this model to the TCCpM data, as well as to the total number of recovery cases, using then the *builtin* function **FindFit** to find the best fitting parameters T_EE_, *X*_EC_, and *σ* as a function of time. This was done by starting with data from only a few days after the first day of the outbreak and then calculating the best fitting parameters for the data of that time frame. After that, the data is updated on a daily basis and a new set of best fitting parameters is found for each day. After enough days, we have a graphic representation of how the fitting parameters changed as new data became available. At first, the fitting parameters vary in an oscillatory way with time, becoming, eventually, stable. When their values stabilise, enough data has been collected to provide more precise fitting parameters, that are now capable of better describing the TCCpM curve by with Eq. (2).

Figs. 4(a-c) give the time evolution of the fitting parameters of Eq. (2) for Hubei for the Total Confirmed and Recovered Cases (TCC and TRC), as well as the fitting curve for TCCpM in (d). From Figs. 4(a-c) we can see that the TCCpM parameters have already stabilised, while the recovery parameters stopped oscillating and are developing in a linear fashion. This is expected as the number of recoveries is still growing. The stability point of the TRC parameters, however, is not expected to be the same as the TCCpM, the difference being accounted by the death cases. Fig. 4(a), however, shows that the amplitude of both cases will become constant at close values, indicating a relatively low mortality rate. In Fig. (b), the difference between the two curves indicates the mean time between contagion and healing, almost 20 days.

**Figure 4:**
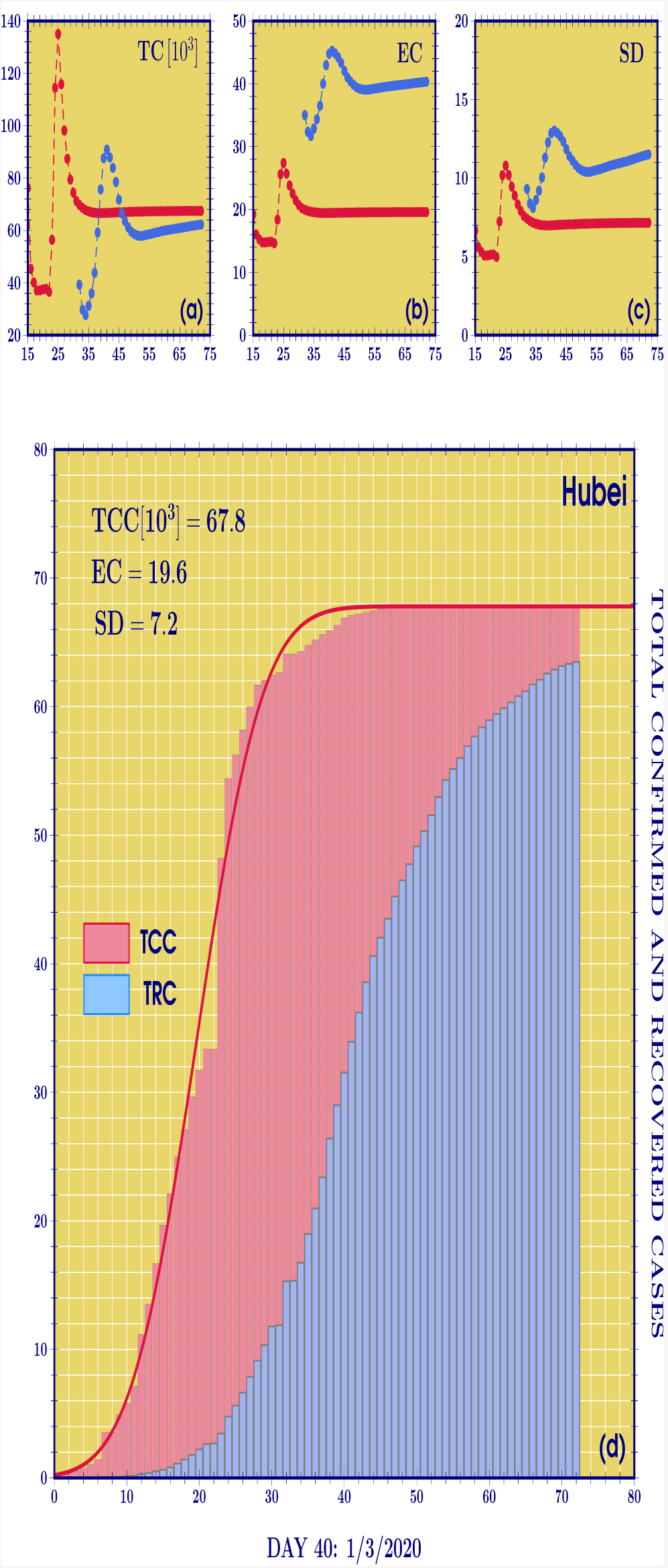
HUBEI. The time evolution of modelling parameters for Hubei: Total Confirmed and Recovered Cases per Million, TCCpM and TRCpM, in (*a*), Epidemic Center, EC, in (*b*), Standard Deviation, SD, in (*c*). Each point in each plot is obtained by finding the best fitting parameters from the TCCpM and TRCpM data up to the time corresponding to that point. When the curves stop varying and become constant with time, the constant values indicate the appropriate fitting parameters. The curve modelling the TCCpM and TRCpM in Hubei is plotted in (d) by using the last point in each of the top graphics. The vertical bars indicate the real data.

Finally, Fig. 5 shows the time evolution of the Epidemic Center for the 10 European countries. Many countries seem to have started the stabilisation phase, which would allow more trustworthy forecasts. A stabilisation of the epidemic center between day 60 and day 70 is seen for Switzerland, Italy, Austria, and probably France. Around day 70, we find the stabilisation of Spain, Netherlands, Portugal, and probably Germany. In contrast, the UK and Belgium curves seem still to be oscillating, and for the Belgium curve its oscillation amplitude seems to be increasing, making forecasts more difficult.

**Figure 5:**
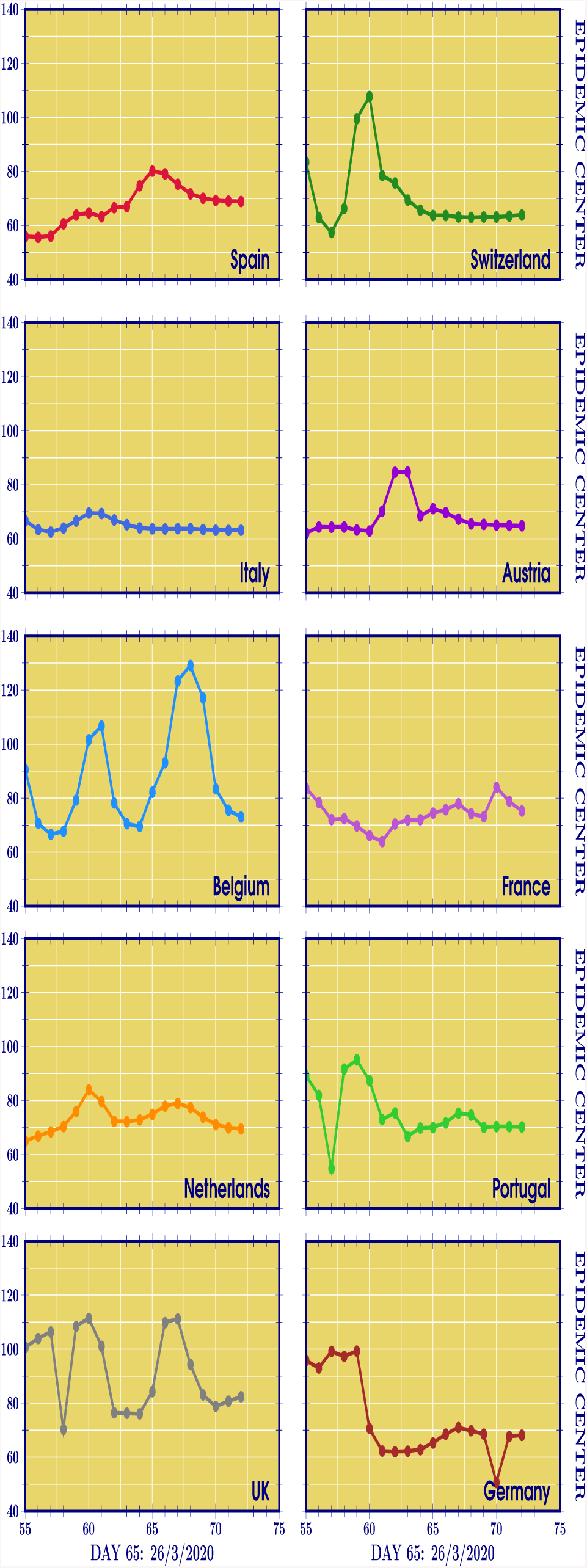
EUROPE. The time evolution of the EC parameter for European countries in analysis. A stabilisation of the epidemic center between day 60 and day 70 is seen for Switzerland, Italy, Austria, and probably France. Around day 70, we find the stabilization of Spain, Netherlands, Portugal, and probably Germany. Belgium and the UK still are in their oscillation phase.

## IV. Italian regions

When analysing the outbreak in a country it is very important to observe that different areas can show very different developments. Consequently, it would be more appropriate to talk of a mean prediction for the country.

Specifically for Italy, the virus seems to be more widespread in the Lombardia region, which is also the highest populated region in the country, see Fig. 6. Looking at Fig. 7(a), we can see that the Italian regions have already reached a *horizontal development* stage, with the *α*-factors of Lombardia, Emilia-Romagna, and Veneto starting to drop, while Piemonte’s curve is flattening out, indicating a linear growth of the spreading rate. In the same figure, we also find the curve for the rest of Italy where the maximum of *α* factor is below 50, a factor five times smaller than the Lombardia maximum. In Fig. 7(b), we find a more detailed look at the *α*-factor, ranging from 0 to 500 TCCpM. We can see that, after the range from 0 to 200, Piemonte and Lombardia’s *α*-factors start to increase compared to the other Italian regions. In this case, with the same measures adopted, the difference in the future spreading rate comes from the recovered over confirmed ratio. This ratio is plotted in Fig. 8. We can see the recovering rate increasing, specially for Lombardia, with a more than 25% recovery rate at day 72 (2 April). The Piemonte region started with a very flat and close to zero recovery rate. This means that the measures for this region were applied when the outbreak was in its initial phase. This explains why, notwithstanding the high *α* factor near 500 TCCpM, Piemonte stabilises below Lombardia and Emilia-Rommagna, where the measures were taken when the outbreak was already ongoing.

**Figure 6:**
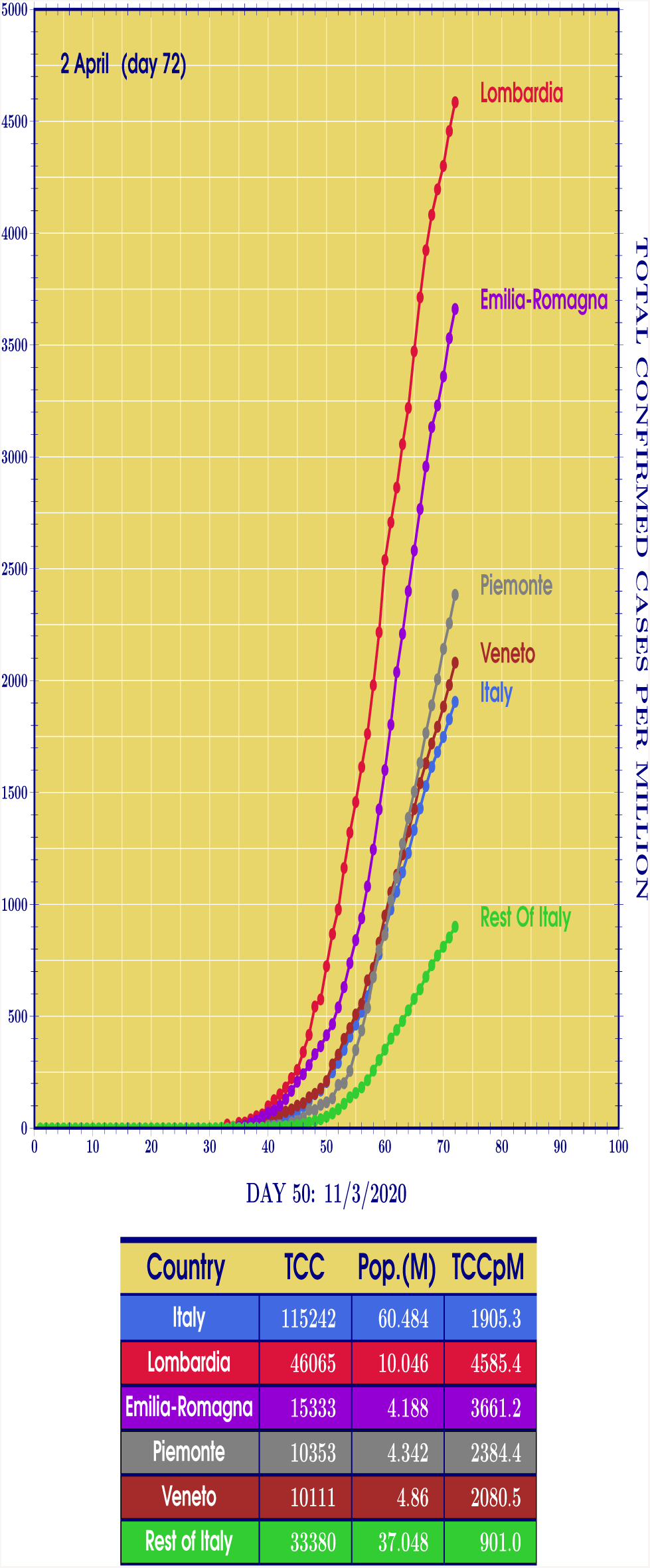
ITALIAN REGIONS. Curves of the Total Confirmed Cases per Mil-lion (TCCpM) of residents for Lombardia, Emilia-Romagna, Piemonte, and Veneto at the 2nd of April (day 72). For the Lombardia and Emilia-Romagna curves, we clearly see an increase in the TCCpM much greater than the ones for the Piemonte and Veneto regions. As a consequence of it, keeping in mind that these four regions represent 38, 7% of the Italy population, the curve for the rest of Italy shows an almost linear behaviour.

**Figure 7:**
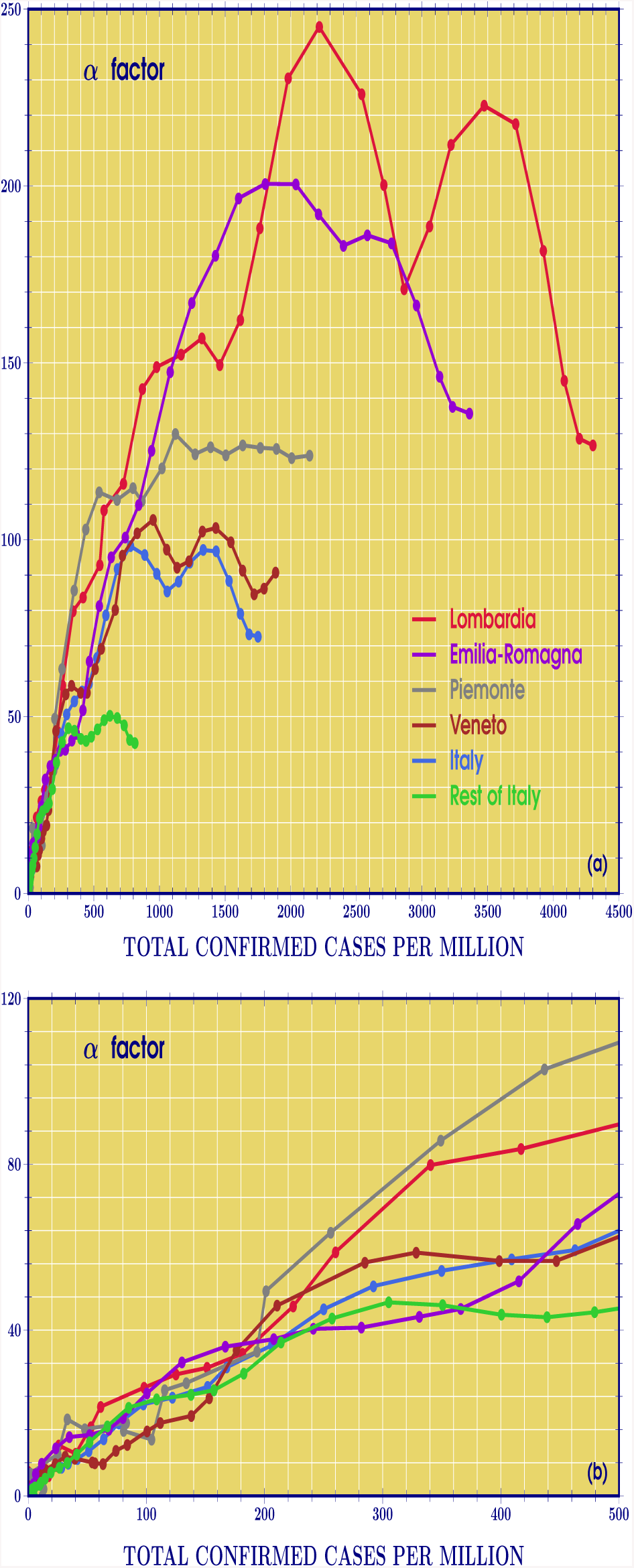
ITALIAN REGIONS. The *α*-factor, representing the spreading rate of COVID-19, is plotted in (a) for Lombardia, Emilia-Romagna, Piemonte, and Veneto as a function of the TCCpM. From the plots it is clear that the four regions reached the maximum of the *α*-factor. The bottom figure (b) allows a more detailed look at the *α*-factor from 0 to 500 TCCpM. We can see that, in the range from 0 to 200, the contagion rate for the Italian regions is approximately the same. After such an initial range, it becomes evident that Piemonte and Lombardia start to increase compared to the other Italian regions.

**Figure 8:**
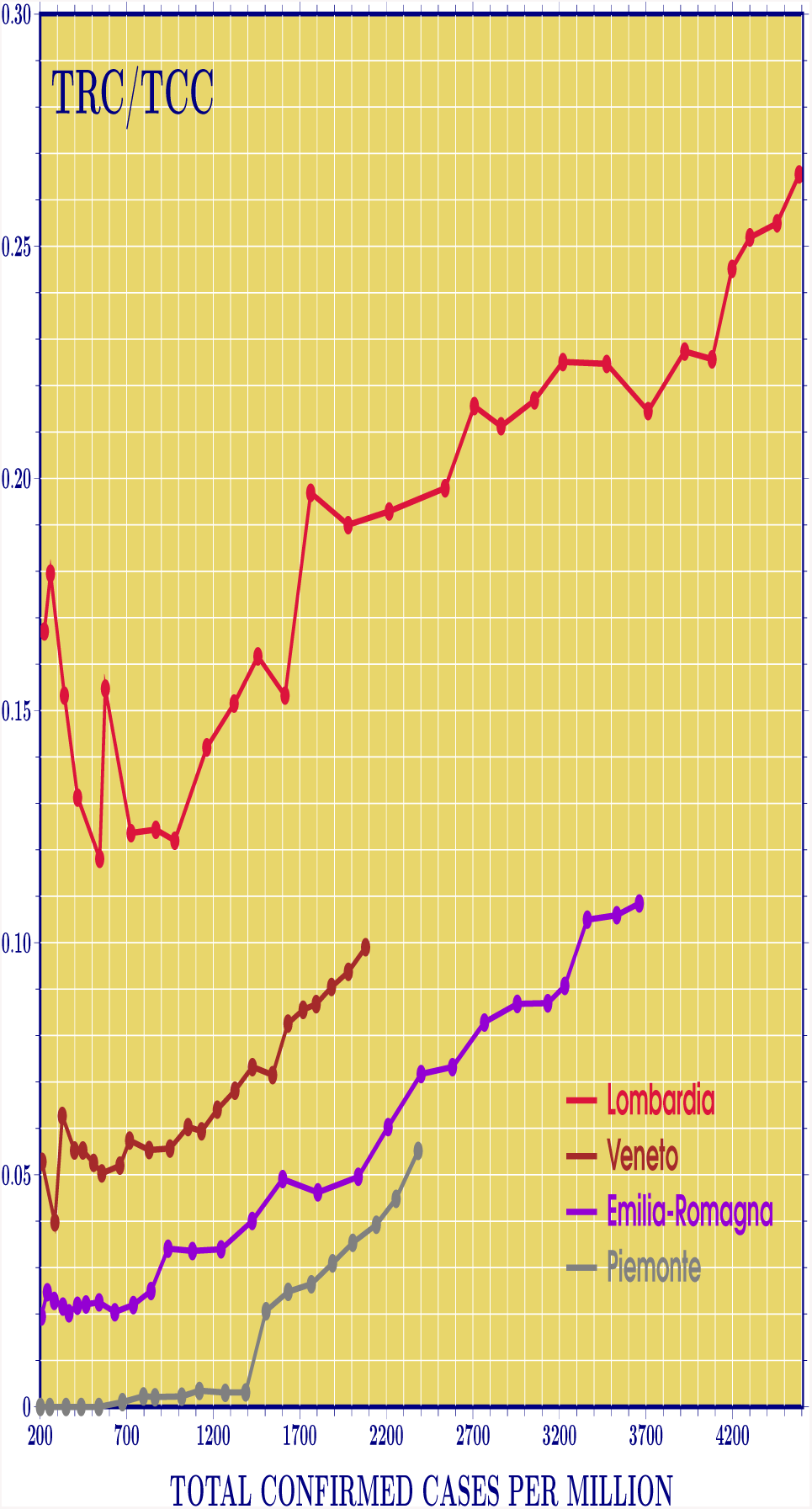
ITALIAN REGIONS. The TRCpM over TCCpM ratio is plotted for Lombardia, Emilia-Romagna, Piemonte, and Veneto. The plots help to complete the analysis of the *α*-factor done in Fig. 8. The plots clearly show that for a given TCCpM point, the Piemonte region has a very low ratio of recovered over confirmed cases compared to the other regions. This means that, according to our interpretation, the measuares to contain the outbreak in Piemonte was taken when it was in its initial phase.

## V. South Ameriica

In the South American continent, while Brazil has the largest number of total confirmed cases, Ecuador and Chile have the greatest number of TCCpM, having nearly 5 times more cases per million residents than Brazil. Ecuador and Chile have shown the most concerning TCCpM rates and, even though their TCCpM is still smaller than that of the European countries, their cases are analogous to the case of Switzerland, with a more widespread infection, see Fig. 9. Similarly, Uruguay has only 3.5 million residents, but the third largest number of TCCpM of the continent.

**Figure 9:**
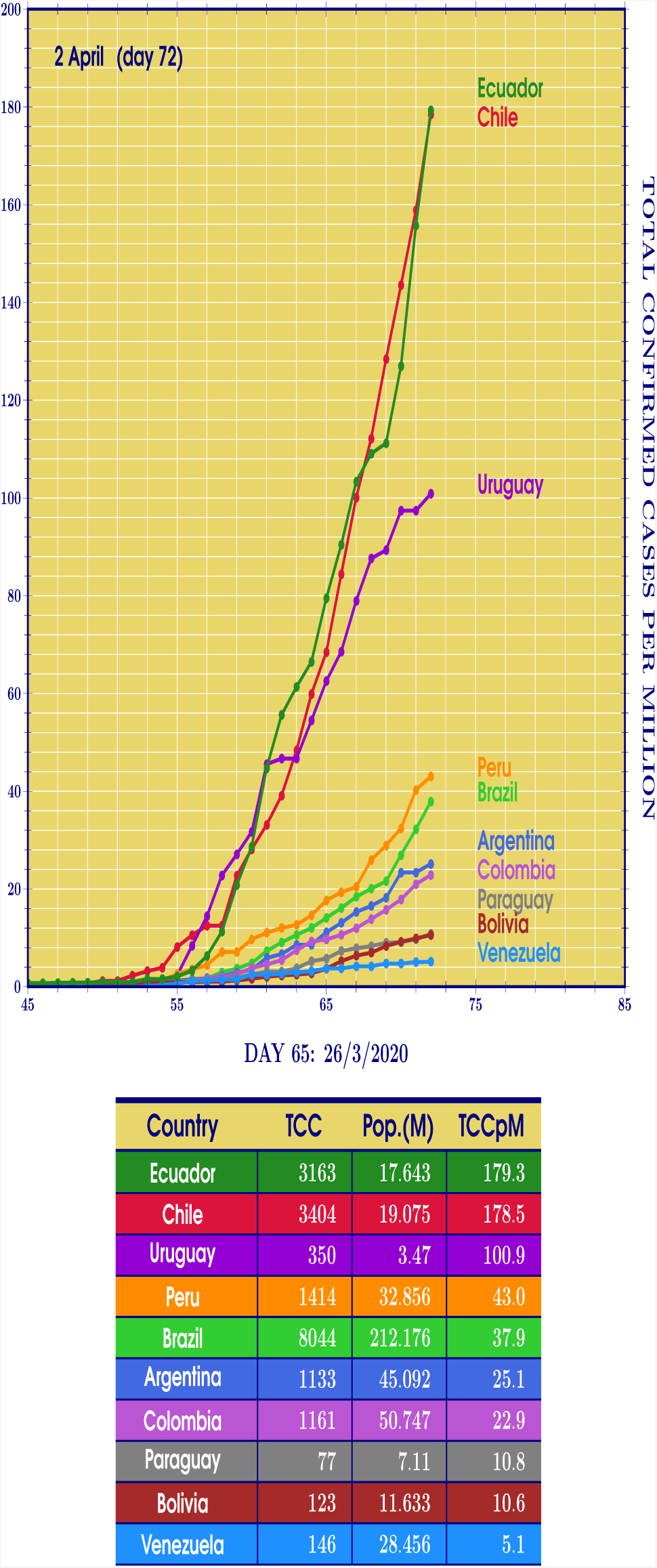
SOUTH-AMERICA. Curves of the Total Confirmed Cases per Million (TCCpM) of residents for Ecuador, Chile, Uruguay, Peru,Brazil, Argentina, Colombia, Paraguay, Bolivia, and Venezuela. From the plots, Ecuador, Chile, and Uruguay show a sharp increase in the TCCpM. A second group (Peru, Brazil, Argentina, Colombia) is characterised by a lower increase compared to the first group. Below, we find Paraguay and Bolivia. Finally, an almost linear increase of TCCpM for Venezuela.

The South American countries in general, see Fig. 10, still show signs of an increasing spreading rate with Ecuador and Chile displaying the largest increase. Fig. 11 compares the early development of the *α*-factors of South American and European countries. The magnitude of the spreading rate is the same for both continents, but while European curves seem to develop in greater agreement amongst each other, in South America, Ecuador, Chile and Uruguay single themselves out very quickly. From Fig. 11, we can see that, as the disease starts to spread in a country, its spreading rate is similar to that of any other country. As more and more people are infected, however, this development becomes more discrepant. These differences might be the result of several different factors such as population density and cultural habits, as well as the earliness and effectiveness of the local authorities health measures.

**Figure 10:**
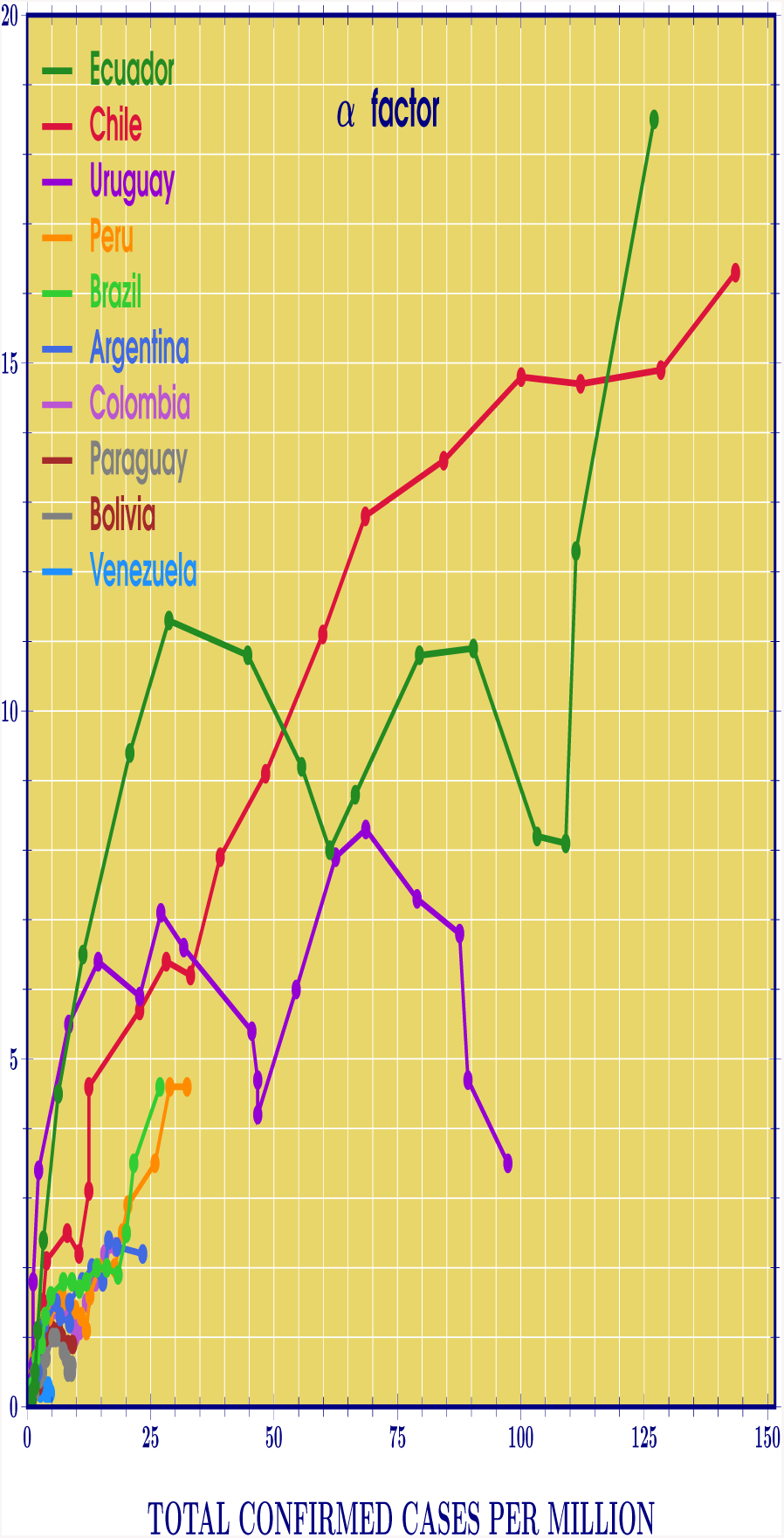
SOUTH-AMERICA. The *α*-factor, representing the spreading rate of COVID-19, is plotted for the South American countries as a function of the TCCpM. The Ecuador and Chile curves display a significant growth, Uruguay seems to have reached the maximum region. The other South American countries follow a very similar path in the of the disease.

**Figure 11:**
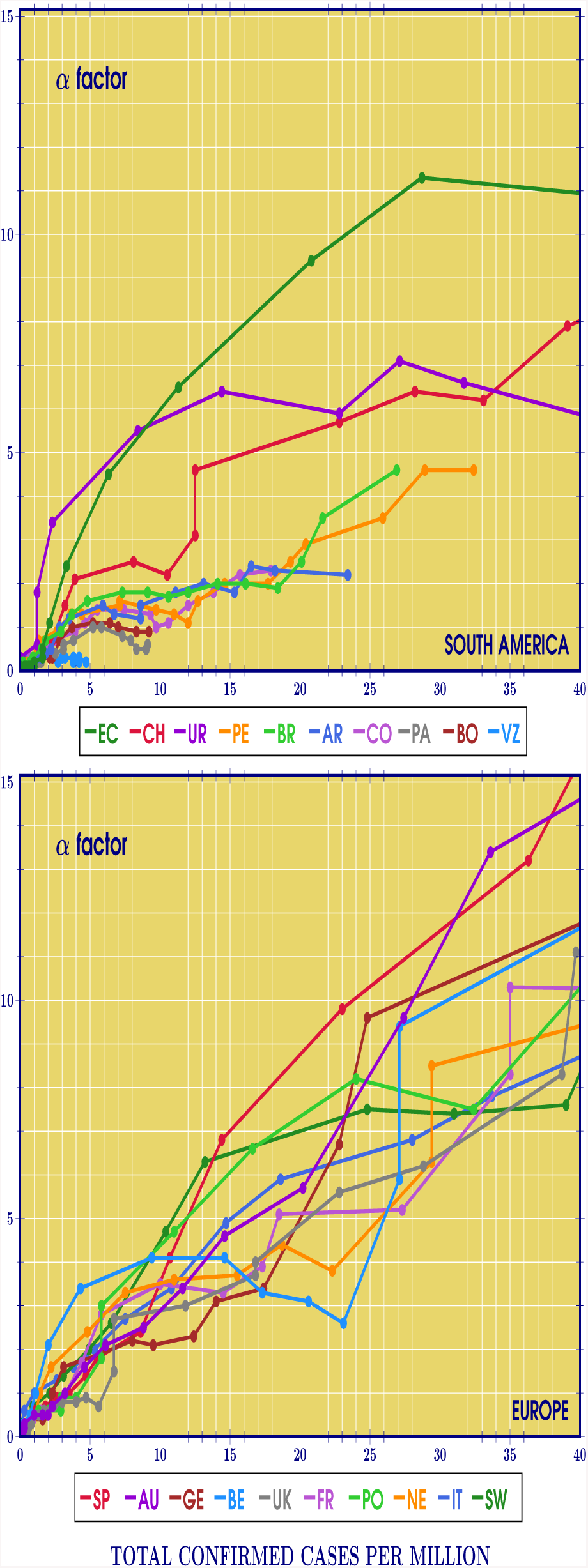
SOUTH-AMERICA/EUROPE. Comparison between the early development of Covid-19 outbreak in South America and Europe. The lower *α* factor in the initial outbreak of the majority of the South American countries with respect to the European ones suggests that, by anticipating the protective measures in the initial stage of the disease, the majority of the South American countries have been successful in preventing an in-creased spreading of the outbreak in comparison to Europe.

For Brazil, the local health measures were taken timely, starting around day 52 (13 March when the TCCpM was only of 0.7). After a week, the Brazilian *α* factor stabilised around 2 and remained at this constant value (indicating a linear spreading rate) until day 67 (28 March). At the end of March, from the Brazilian data we find

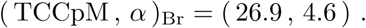

In comparison to Italy, Switzerland, and Spain

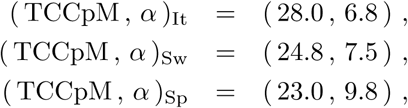

Brazil shows a lower spreading rate with respect to the European countries. The Brazilian as well as the other South American countries data will be constantly updated in the website [8].

## VI. Conclusions

We have studied the data on the COVID-19 pandemic for 22 countries and 4 Italian regions, analysing how fast it spreads as more people are contaminated. We have also analysed how to model the total number of confirmed cases, and how the parameters of the model change as more information on the number of affected people becomes available. We can see that as the infection spreads, these parameters oscillate with time until they become stable. The values at which they stabilise then gives us a more trustworthy basis on which to simulate the outcome of the pandemic.

In regard to the spreading rate, we have studied how it changes as the number of confirmed cases increase. At first, all countries’ rates develop in a similar fashion, which changes as more people are contaminated. This could reflect the different responses to the pandemic given by different countries. So far, only China and South Korea have dropped this rate to zero, showing close to none daily new cases. South Korea in particular, is a very interesting case, having dropped this rate to zero early on in the pandemic development. In South America, Venezuela and Paraguay also show signs of an early decrease in the spreading rate of COVID-19.

The modelling of the Total Confirmed Cases per Million (TCCpM) residents curve was done by the asymmetrical integral of a Normal Distribution [9]. By fitting the TCCpM data to this model in progressively larger ensembles, we were able to study how the model’s parameters change with time. We can see in the modelling of the province of Hubei in China that the parameters values oscillate for some time until stabilising as constants. These constants can then be used in the model to simulate the development of the pandemic. Italy’s parameters are already showing signs of stabilisation, with a forecast that the TCCpM will reach its peak by the end of April. Other European countries have also shown such signs, with the exception of Belgium and the UK, which still show a strong oscillatory pattern.

We hope, the analysis presented in this paper, based on the joint use of the *α* factor, the graphical representations of the oscillating fitting parameters, and the recovery cases over confirmed cases ratio, will contribute to the study of the effectiveness of the methods employed by different local authorities in containing epidemics, as well as to the study of the readiness with which such methods must be activated. By analysing more closely the Italian case, looking at the spreading of COVID-19 in different Italian regions, we have shown how, even inside the borders of a country, epidemics may develop in different ways. The analysis done can be helpful in aiding national health authorities in planning and spending resources appropriately in different regions under their jurisdiction.

The model’s fitting parameters, see Fig, 4 and 5, were obtained by using a normal probability density function. This allows to identify the daily peak with the epidemic center. Due to the fact that after the daily peak, it is possible to have an asymmetric decrease in the number of daily confirmed cases a more real analysis should be done by using skew normal probability density functions. This will be the objective of our future investigations.

## Data Availability

http://www.ime.unicamp.br/∼deleo/CoVid19.html

http://www.ime.unicamp.br/~deleo/CoVid19.html

## Final acknowledgements

The authors would like to express their special thanks to Dr. Rita K. Kraus for her constructive scientific suggestions during the planning and development of this research work, to Fabio Valente for valuable conversations on the timing of the measures taken by the Italian Government, and, last but not least, to Piero Trinchera for his daily assistance on the Italian health data collection.

Our heartfelt thanks go to physicians and nurses who, during the last months, have fought on the front line against this invisible enemy. Their unbelievable bravery, constant dedication, and continuous effort gave to the world a hope to win this tough battle.

---

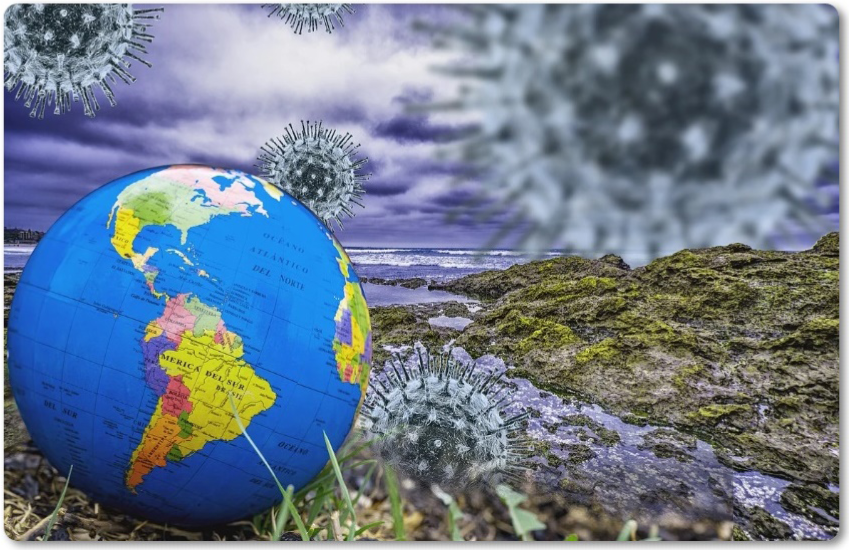

